# Dynamic analysis and evaluation of asymptomatic infection in the spread of COVID-19

**DOI:** 10.1101/2021.12.07.21267442

**Authors:** Chuanqing Xu, Zonghao Zhang, Xiaotong Huang, Jingan Cui

## Abstract

COVID-19 has spread worldwide for nearly two years. Many countries have experienced repeated epidemics, that is, after the epidemic has been controlled for a period of time, the number of new cases per day is low, and the outbreak will occur again a few months later. In order to study the relationship between this low level of infection and the number of asymptomatic infections, and to evaluate the role of asymptomatic infections in the development of the epidemic, we have established an improved infectious disease dynamics model that can be used to evaluate the spread of the COVID-19 epidemic, and fitted the epidemic data in the three flat periods in England. According to the obtained parameters, according to the calculation of the model, the proportion of asymptomatic infections in these three flat periods are 41%, 53% and 58% respectively. After the first flat period, the number of daily newly confirmed cases predicted by the model began to increase around July 1, 2020. After more than four months of epidemic spread, it reached a peak on November 12, which is consistent with the actual case situation. Unanimous. After the second flat period, the model predicts that the number of new confirmed cases per day will increase from about May 7, 2021, and after about 73 days of epidemic development, it will reach a peak on July 20, showing the overall trend of the epidemic. In the above, the predicted results of the model are consistent with the actual cases. After the third flat period, the number of daily newly diagnosed cases predicted by the model began to increase around December 1, 2021, and reached a peak in December, and the number of cases will drop to a very low level after May 2022. According to our research results, due to the large number of asymptomatic infections, the spread of the epidemic is not easy to stop completely in a short time. However, when the epidemic enters a period of flat time, nucleic acid testing is performed, and asymptomatic infections are isolated at home for 14 days (the recovery period of symptomatic infection is about 10 days) may be an option that can be considered to interrupt the transmission of the case.

## Introduction

Since the outbreak of the COVID-19 epidemic at the end of 2019, it has had a serious impact on countries around the world. How to effectively control the spread of the epidemic and restore normal life has become an urgent issue for governments of all countries. As of November 2021, the cumulative number of confirmed COVID-19 patients worldwide has exceeded 250 million. With the advent of winter, Europe and other regions have begun a new round of outbreaks. There are a large number of asymptomatic infections with a certain degree of infectiousness among the infected persons of COVID-19. Due to the existence of asymptomatic infection, the spread of SARS-CoV-2 is more insidious, which makes the prevention and control of the epidemic more difficult.

Asymptomatic infection, in this article, refers to an infected person who will not show symptoms throughout the infection period. Many clinical studies have shown that asymptomatic infections of COVID-19 are infectious ^[1,2,3,4]^. As it is difficult for asymptomatic infected people to be aware that they have been infected, their social behavior and activities will not be affected in any way, and they will still be active in the susceptible group, which will play a certain role in promoting the spread of the epidemic.

There have been a lot of results in the research on asymptomatic infection of COVID-19. Asymptomatic infections occur between 0 and 73 years old ^[5]^. The asymptomatic proportion of infected persons averaged about 46% ^[6]^. The transmission intensity of asymptomatic infection is about 65.24% of that of symptomatic infection ^[7]^. In addition, some researchers have established a compartmental model that includes asymptomatic infections to study the spread of COVID-19. Ruan et al. established a time-varying COVID-19 transmission compartmental model containing asymptomatic infections, reviewed the development process of the Wuhan epidemic, and found that asymptomatic infections accounted for about 20% ^[8]^. Rahul Subramanian and others established a COVID-19 transmission model including asymptomatic infections, and quantified the asymptomatic infections and transmission in New York City, and the proportion of asymptomatic infections was about 60% ^[9]^. Salihu S. Musa and others established a SEIR COVID-19 infectious disease compartmental model including asymptomatic infections, and analyzed the spreading dynamics of the epidemic in Nigeria ^[10]^. Mohamed Amouch and others proposed a new epidemiological mathematical model for the spread of COVID-19 disease, and simulated the outbreak in Monaco, and found that the asymptomatic proportion of infected persons was 30% ^[11]^.

It has been nearly 2 years since SARS-COV-2 appeared, and various countries including China have experienced repeated epidemics. Since the end of the Wuhan COVID-19 epidemic in June 2020, many small-scale epidemics have occurred in many areas of China, including Sui Fenhe, Mudanjiang, Beijing, Guangzhou, Dalian, Yangzhou, Xiamen, etc. After October 2021, more than 20 cities, including Beijing, have experienced COVID-19 again. China has adopted different epidemic prevention and control measures than most countries, mainly through large-scale nucleic acid testing, temporarily closed and managed areas where the epidemic occurred and communities where cases occurred, and centralized isolation of close contacts, and finally eliminated the epidemic. This also allows China to maintain a nearly epidemic-free state in China, which is very similar to the quantum state in physics. Some countries and regions have also carried out multiple rounds of lockdown or isolation measures to control the further spread of the epidemic, such as Canada, England, Japan, etc. These countries and regions all began to gradually lift the epidemic prevention measures when the epidemic reached a low level, and after a few months, the epidemic rebounded. We tried to estimate the number and proportion of asymptomatic infections during this low-level period through modeling. And further analyzed whether as the asymptomatic infections accumulate, after a period of time, the outbreak of the epidemic will occur again.

How to evaluate the role of asymptomatic infection in the spread of the epidemic, especially in the early stage of the epidemic, is particularly important. If the number of asymptomatic infections can be effectively estimated before the outbreak, the early warning mechanism is activated when asymptomatic infections accumulate to a certain threshold, and certain epidemic prevention and control measures can be taken to delay the spread of the epidemic to a certain extent. This article aims to establish a COVID-19 transmission dynamics model to evaluate the role of asymptomatic infection in the development of the epidemic.

## 1. Research object

Repeated outbreaks have occurred in many countries, including England. This article chooses to study the data of daily new cases in England, and the data comes from the official website of the British government. The epidemic case data in England in the past two years shows such typical characteristics, as shown in Fig.1. From June 1 to July 31, 2020, the daily number of new cases was at a low level, with an average of 434 new confirmed cases per day. This time period is recorded as the first flat period. Subsequently, the epidemic broke out further, with the first and second peaks consecutively appearing, of which the second peak was mainly caused by the Alpha variant. After a period of lockdown and censorship management, the epidemic will again enter a relatively low level from April 1 to May 31, 2021, with an average of 2137 newly diagnosed people per day. This time period is recorded as the second flat period. Since then, the epidemic broke out again. After another lockdown and control measures, from September 1, 2021 to the time of writing, the epidemic was at a relatively low level, with an average of 28,742 newly diagnosed people per day. The number of confirmed cases per day is much higher than the lower levels of the previous two times. This time period is recorded as the third flat period. Therefore, we have established a COVID-19 transmission dynamics compartmental model, and performed a fitting analysis of these three parts of typical data to study the impact of the presence of asymptomatic infections on the recurrence of the epidemic.

**Fig.1.**
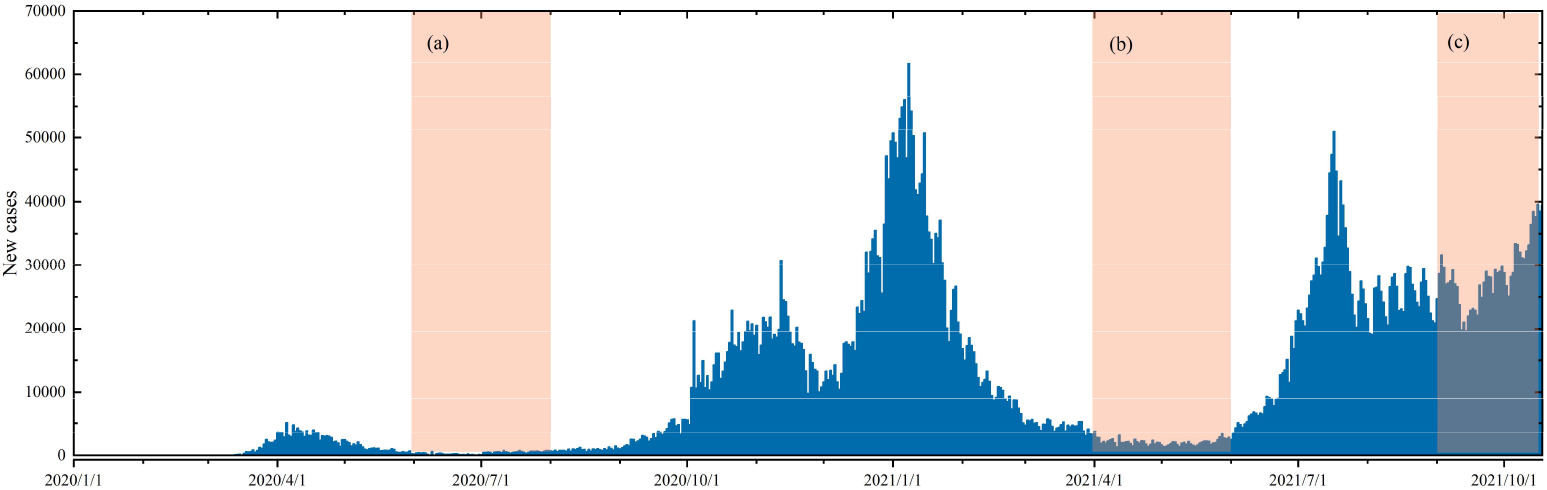
The blue histogram represents the daily new confirmed cases in England. The three orange areas (a), (b), (c) represent the time period of the selected research object. Area (a) represents the first flat period, from June 1 to July 31, 2020. Area (b) represents the second flat period, from April 1 to May 31, 2021. Area (c) represents the third flat period, from September 1, 2021 to the time of writing this article.

## 2. Model establishing

The main ways of transmission of COVID-19 are direct transmission, aerosol transmission and contact transmission. According to whether symptoms appear, COVID-19 infected persons can be divided into asymptomatic infections and symptomatic infections. Symptomatic infections are clinically manifested as fever, dry cough, and fatigue. Among them, severely ill patients will experience organ failure or even death.

Based on the cognition of COVID-19, we have the following assumptions in the model:

1. Due to the long exposure period of COVID-19 patients and the difference in infectiousness, we divide the exposure period into two parts: the pre-exposure period and the post-exposure period. The early period is not infectious, and the later period is infectious.
2. Symptomatic infections are divided into two compartments for testing and non-testing according to the actual situation.
3. Assume that asymptomatic infections can be screened as long as they are tested, and the detected symptomatic infections are completely isolated and no longer transmittable. Assume that asymptomatic infections are not tested.
4. The birth rate and natural death rate at the population level are not considered, and the death rate due to disease of asymptomatic infections is not considered.

Based on the above assumptions, we have established a COVID-19 transmission compartmental model containing asymptomatic infections. The model divides the total population of England into susceptible persons (S), patients in the pre-exposure period (E1), patients in the post-exposure period (E2), symptomatic infections who are tested (I1), and symptomatic infections who are non-tested (I2), asymptomatic infections (A), recovering (R). The kinetic flow chart is shown in Fig.2.

**Fig.2.**
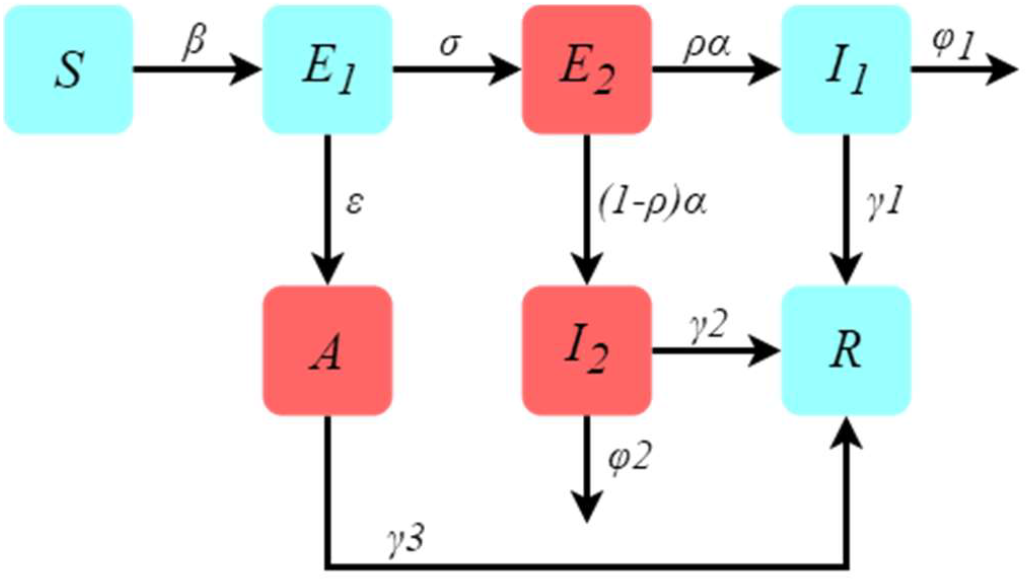
This figure shows the flow chart of the COVID-19 transmission dynamics model containing seven compartments. Among them, the compartments represented by the red box is infectious, and the compartments represented by the blue box is not infectious.

The corresponding propagation dynamics equation is constructed as follows:

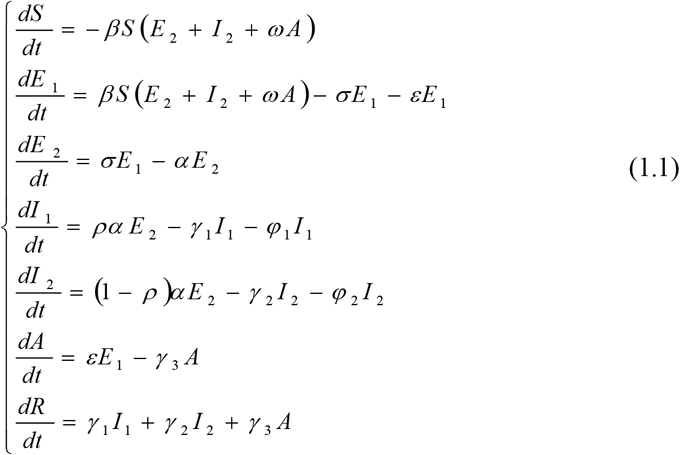

Here, β represents the basic transmission rate. σ represents the conversion rate of patients in the pre-exposure period to patients in the post-exposure period. ε represents the conversion rate from patients in the pre-exposure period to asymptomatic infections, α represents the conversion rate from patients in the post-exposure period to symptomatic infections. ω represents the transmission intensity of asymptomatic infection relative to symptomatic infection. ρ represents the rate of symptomatically infections who are tested. γ_1_ indicates the recovery rate of symptomatic infections tested. γ_2_ indicates the recovery rate of symptomatic infections non-tested. γ_3_ indicates the recovery rate of asymptomatic infections. φ_1_ represents the mortality rate of symptomatic infections tested. φ_2_ is the mortality rate of symptomatic infections non-tested.

### 2.1 The disease-free equilibrium point of the model and the controlled reproduction number

The disease state bins in the system (1.1) are E1, E2, I1, I2, A. In order to calculate the control reproduction number of the system (1.1), we take:

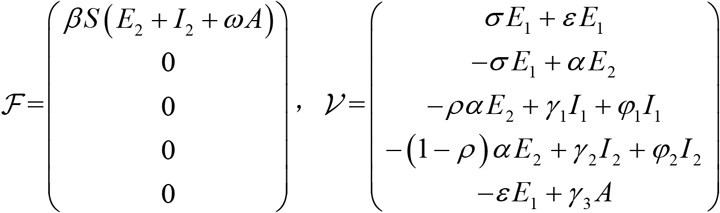

The Jacobian matrices of ℱ and 𝒱 are obtained as:

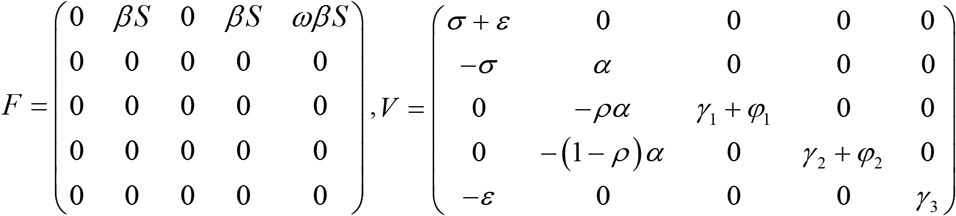

Therefore, at the disease-free equilibrium point *P* ^0^ = (*S*_0_ 0 0 0 0 0 *R*_0_), there is:

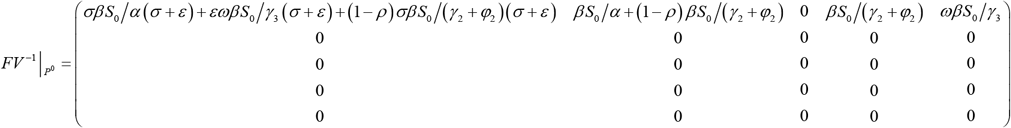

Therefore, the controlled reproduction number is:

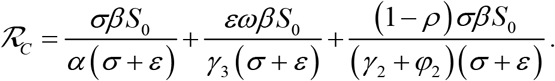

The controlled regeneration number in the system (1.1) has biological significance. The first item indicates that a single patient in the pre-exposure period enters the compartment of the post-exposure period according to the proportion of 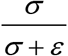, which can cause new infections of *β S*_0_ units within a unit time, and the duration of the post-exposure period is 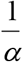; The second item indicates that a single patient in the pre-exposure period enters compartment of asymptomatic infections according to the proportion of 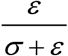, which can cause new infections of *ωβ S*_0_ unit within a unit time, and the duration of the asymptomatic infections period is 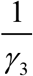; The third item indicates that a single patient in the pre-exposure period enters compartment of symptomatic infections non-tested according to the proportion of 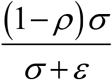, which can cause new infections in unit *β S*_0_ within a unit time, and the duration of the symptomatic infections non-tested is 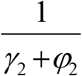. That is, the controlled reproduction number is the weighted sum of the new infections caused by a single patient in the post-exposure period, a single asymptomatic infection, and a single non-tested symptomatic infection during their respective infection periods.

### 2.2 Results and analysis of data simulation

Based on the system (1.1), we used the nonlinear least squares method to simulate the cumulative number of new cases detected during the first flat period in England. The initial value is selected as:S(0)= 55699400, E_1_(0)= 15000, E_2_(0)=300, I_1_(0)=294, I_2_(0)=300, A(0)=300. The simulation results are shown in Fig.3. It can be seen from Fig.3B that the absolute error shows a good agreement between the original data and the simulation results, indicating that the simulation effect is very good.

**Fig.3.**
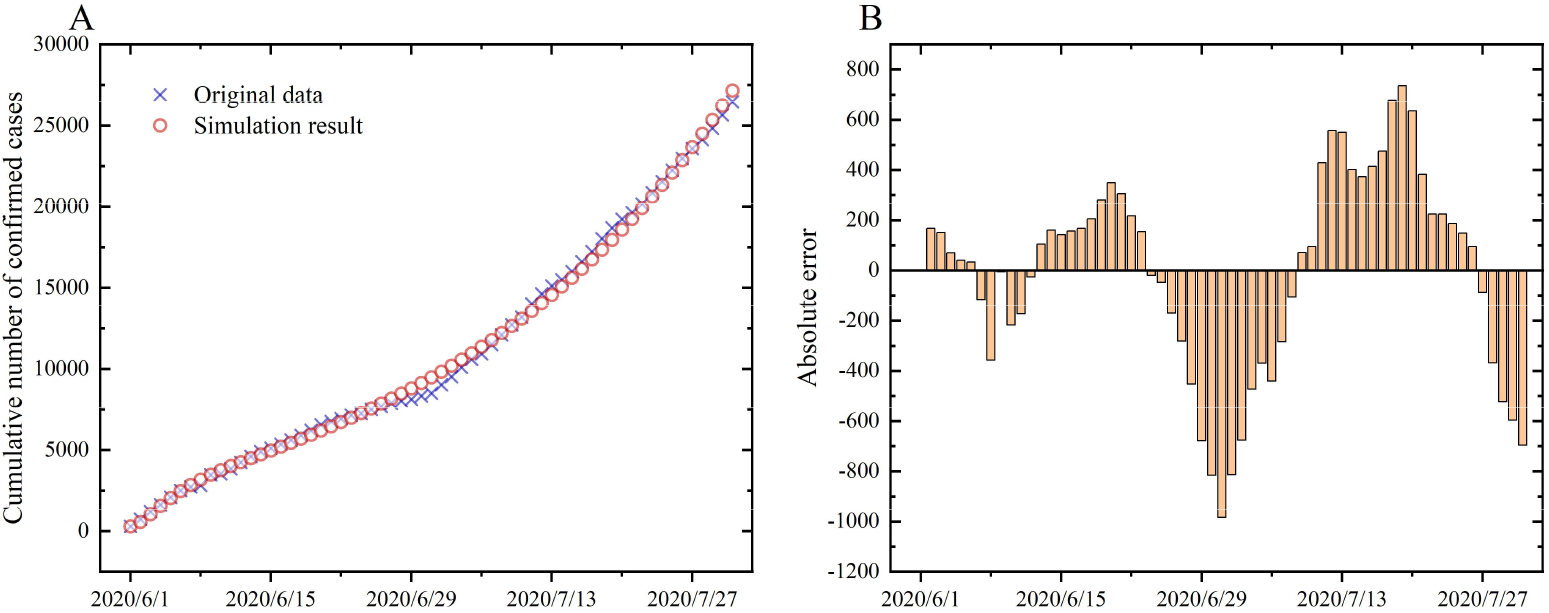
A shows the fitting situation of the cumulative confirmed cases in England from June 1 to July 31, 2020. The blue cross represents the original data, and the red circle represents the fitting result. B shows the absolute error between the original data and the fitting result. The fluctuation of the absolute error between positive and negative indicates that this is a good fit.

Some fitting parameter values are shown in Tab.1. Fittingly, the proportion of the number of symptomatic infections who received testing to the total number of symptomatic infections is ρ=0.3955, which means that during this period in England, the proportion of symptomatic infections being tested was only 39.55%, which is 60.45% of symptomatic infections did not receive nucleic acid testing. The recovery rates of symptomatic infections non-tested and asymptomatic infections are γ_2_=0.0984 and γ_3_=0.0974, respectively, indicating that the recovery period of symptomatic infections non-tested and asymptomatic infections are roughly the same, both about 10 days. The conversion rate from the pre-exposure period to the asymptomatic infections obtained by the fitting is ε=0.1473, which indicates that it takes about 7 days for a patient in the pre-exposure period to develop into an asymptomatic infection.

**Tab.1.**
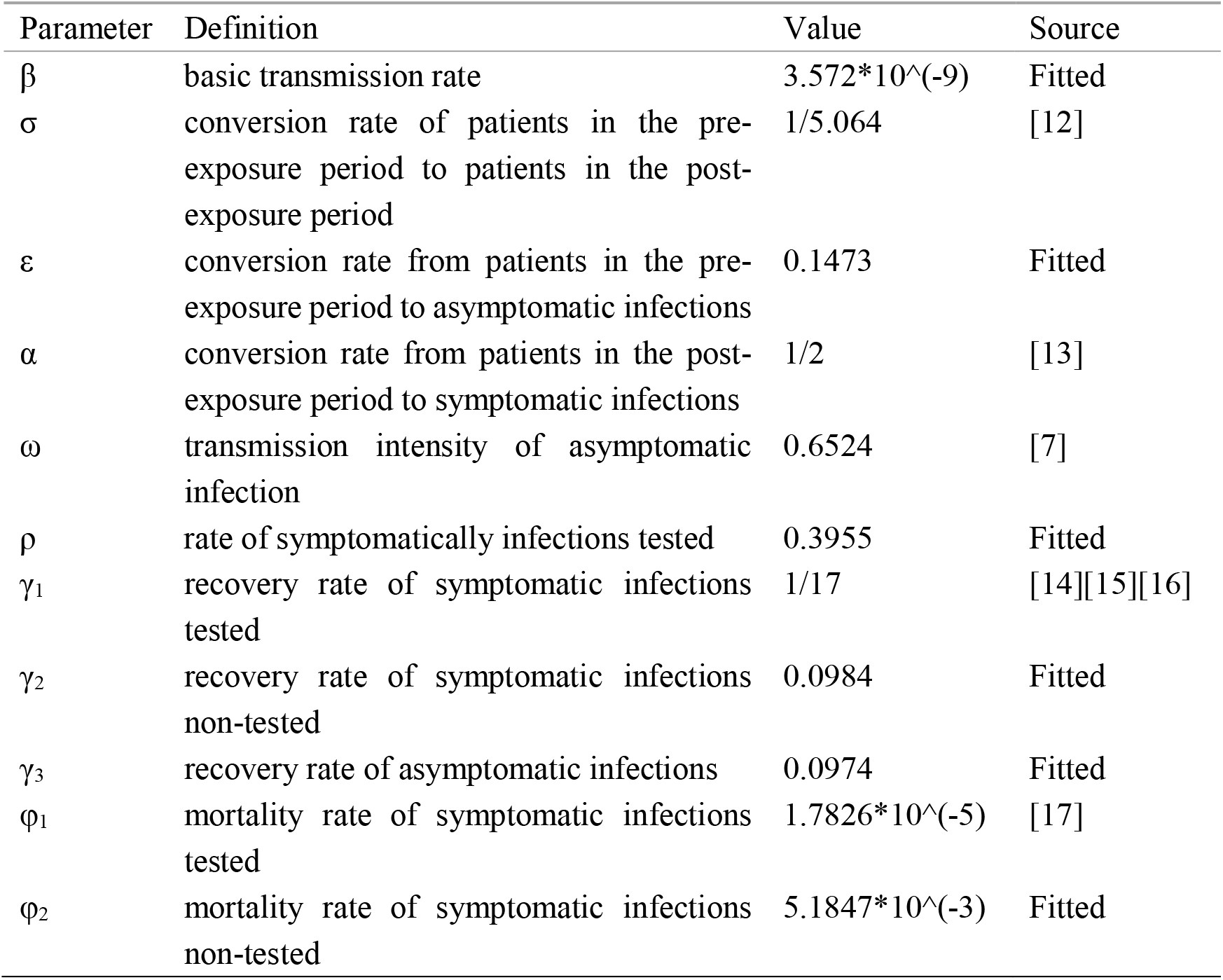
Fitted parameter results during the first flat period

| Parameter | Definition | Value | Source |
| --- | --- | --- | --- |
| $\beta$ | basic transmission rate | $3.572 \times 10^{-9}$ | Fitted |
| $\sigma$ | conversion rate of patients in the pre-exposure period to patients in the post-exposure period | 1/5.064 | [12] |
| $\varepsilon$ | conversion rate from patients in the pre-exposure period to asymptomatic infections | 0.1473 | Fitted |
| $\alpha$ | conversion rate from patients in the post-exposure period to symptomatic infections | 1/2 | [13] |
| $\omega$ | transmission intensity of asymptomatic infection | 0.6524 | [7] |
| $\rho$ | rate of symptomatically infections tested | 0.3955 | Fitted |
| $\gamma_1$ | recovery rate of symptomatic infections tested | 1/17 | [14][15][16] |
| $\gamma_2$ | recovery rate of symptomatic infections non-tested | 0.0984 | Fitted |
| $\gamma_3$ | recovery rate of asymptomatic infections | 0.0974 | Fitted |
| $\varphi_1$ | mortality rate of symptomatic infections tested | $1.7826 \times 10^{-5}$ | [17] |
| $\varphi_2$ | mortality rate of symptomatic infections non-tested | $5.1847 \times 10^{-3}$ | Fitted |

In addition, we used the parameters obtained by the fitting to make predictions, as shown in Fig.4. From the figure, we can see that the number of daily newly confirmed cases predicted by the model began to increase around July 1, 2020, and after more than four months of epidemic spread, it reached a peak on November 12, 2020. In addition to the fact that the number of newly diagnosed daily in the later period is lower than the real situation, the prediction results obtained by the model are more in line with the overall trend of the actual data.

**Fig.4.**
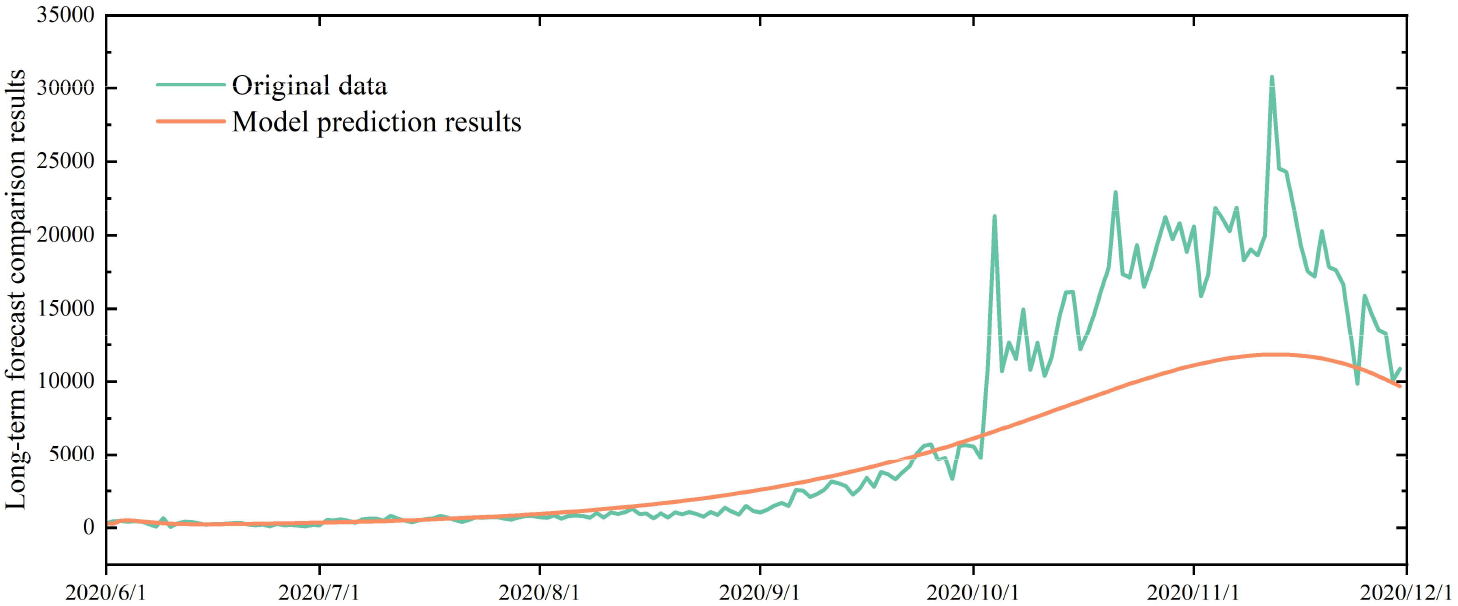
This figure shows the comparison between the model prediction results and the original data. The prediction results of the model are basically consistent with the actual development of the epidemic.

### 2.3 Simulation results and analysis

#### 2.3.1 The impact of the number of asymptomatic infections on the spread of the epidemic

We discussed the impact of the parameter ε_1_ on the spread of the epidemic. Take ε_1_=10%, 15%, and 20% for re-prediction, and the results are shown in Fig.5. As ε increases, the cumulative amount of asymptomatic infections gradually increases, and the cumulative amount of symptomatic infections gradually decreases. Although the total cumulative amount of infected people has increased, this increase is not very obvious.

**Fig.5.**
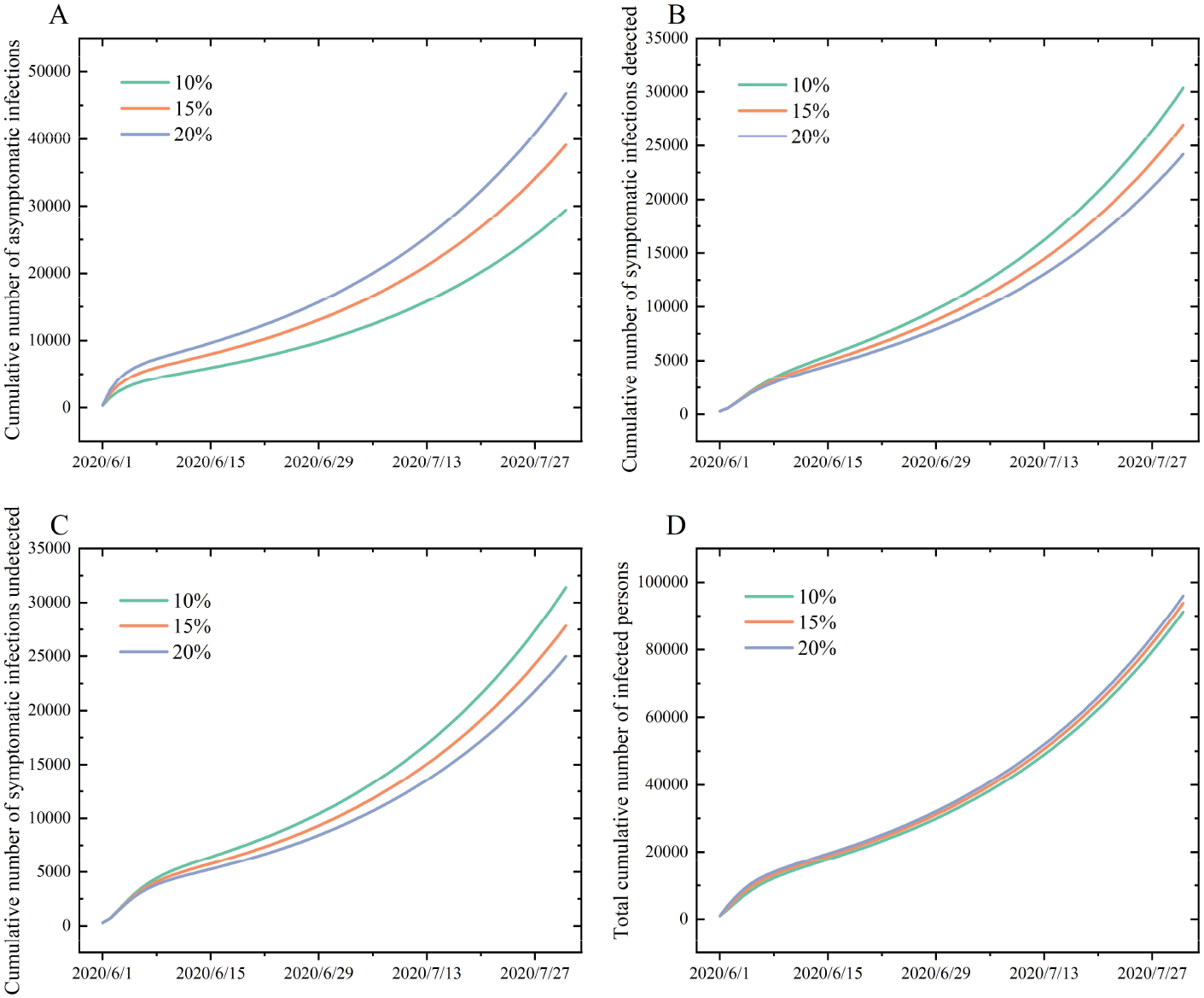
A represents the change in the cumulative amount of asymptomatic infections as ε increases. B represents the change in the cumulative amount of detected symptomatic infections as ε increases. C represents the cumulative amount of undetected symptomatic infections as ε increases. D represents the change of the cumulative amount of total infected persons as ε increases.

In addition, we also analyzed the phase diagrams of asymptomatic infections and symptomatic infections tested over time, as shown in Fig.6B. It can be seen that there is a very strong correlation between asymptomatic infections and symptomatic infections tested.

**Fig.6.**
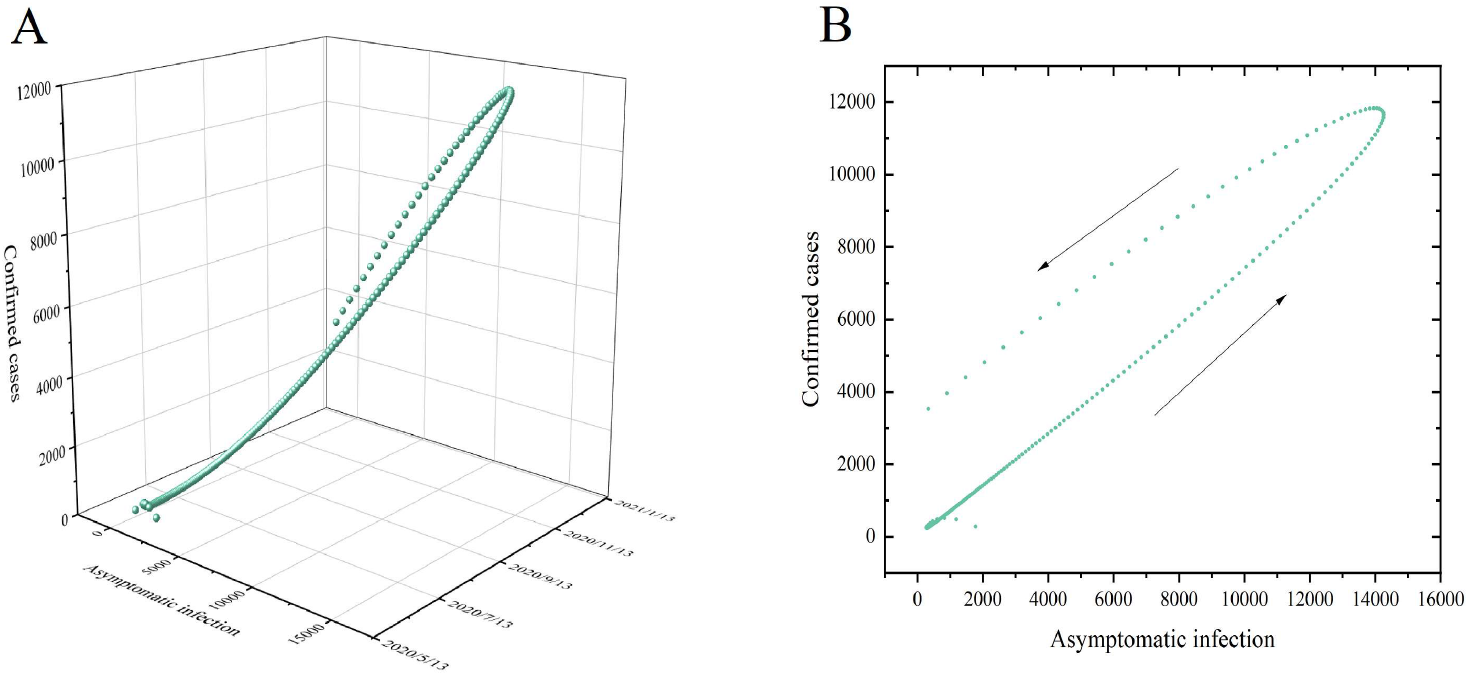
A represents a three-dimensional point diagram of asymptomatic infections and confirmed cases with respect to time. Each sphere represents the value of asymptomatic infections and confirmed cases in the same day. B represents a dynamic phase diagram of asymptomatic infection and confirmed cases with respect to time changes. The direction pointed by the arrow is the direction of time growth.

#### 2.3.2 The impact of the rate of symptomatic infections being tested on the epidemic

In actual circumstances, as the number of newly diagnosed patients increases every day, certain prevention and control measures will inevitably be taken to delay the spread of the epidemic. The most direct and effective prevention and control measure is to increase detection efforts. The increase in detection intensity directly leads to the increase of the parameter ρ, so we will explore the impact of this parameter on the spread of the epidemic. Re-predicted the situations when ρ = 20%, 40%, 60%, and 80% respectively, and the results are shown in Fig.7.

**Fig.7.**
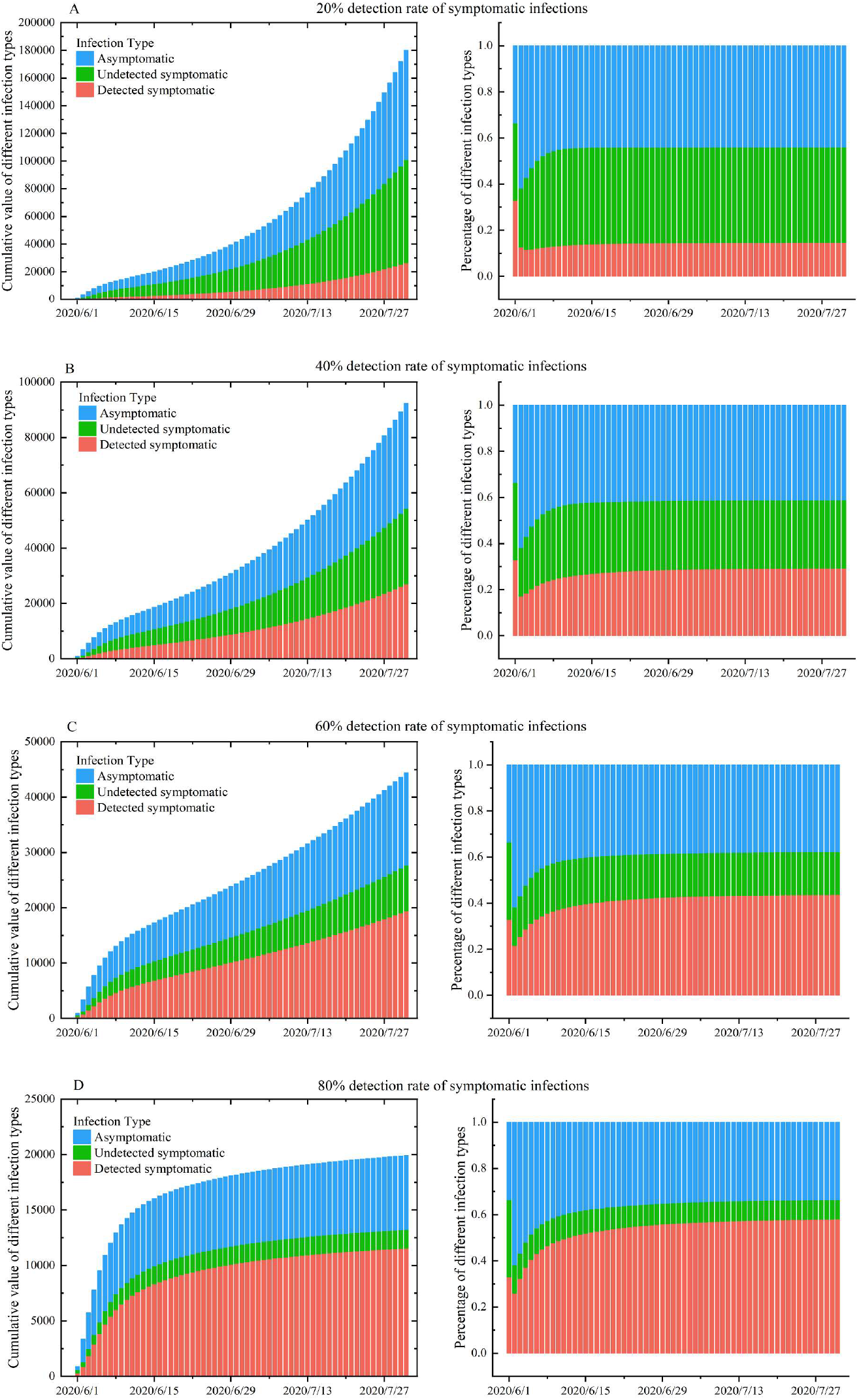
This figure shows the changes in the forecast of the epidemic data in England from June 1 to July 31, 2020, as the detection rate changes. Figures A, B, C, and D show the predictions when the detection rate is 20%, 40%, 60%, and 80%, respectively. The left graph in each sub-graph shows the cumulative value of different types of infected persons, and the right graph shows the proportion of different types of infected persons. Among the types of infected persons, orange indicates symptomatic infections that have been detected, green indicates symptomatic infections that have not been detected, and blue indicates asymptomatic infections.

The proportion of symptomatic infections tested by model fitting is 39.55%. Consider using 40% instead of the result of the parameter ρ obtained by fitting, and the remaining parameters remain unchanged. Through numerical simulation, the cumulative number of cases and their proportions of symptomatic infections tested, symptomatic infections non-tested, and asymptomatic infections are obtained, as shown in Fig.7B. During this time period, asymptomatic infections accounted for approximately 41% of the total infections.

From the results, it can be seen that with the increase of ρ, the cumulative amount of the three different types of infections has decreased significantly. From the perspective of proportion, with the increase of ρ, the proportion of symptomatic infections tested has decreased significantly, and the proportion of symptomatic infections non-tested has increased significantly. In addition, the proportion of asymptomatic infections has declined to a certain extent, but this decline is not significant enough. With the increase in the number of symptomatic infections tested, the role of symptomatic infections in the spread of the epidemic has gradually decreased, which is also the direct reason for the sharp drop in the total number of infected people. However, because asymptomatic infections are not tested, although the transmission intensity is lower than that of symptomatic infections, it will still cause a certain amount of transmission. From the results, it can be seen that even if the proportion of symptomatic infections participating in the test reaches 80%, asymptomatic infections still account for 33%.

### 2.4 Data simulation and analysis in the second flat period

In March 2021, patients infected with the Delta variant appeared for the first time in England ^[18]^. Infected persons with Delta variants are more infectious, have a higher viral load, and have a shorter exposure period ^[19]^. According to current research, the exposure period of patients infected with the Delta variant has been shortened to 4.4 days ^[20]^, and the basic transmission rate has increased by 60% ^[21]^. From April 1 to May 31, 2021, the epidemic in England once again entered a relatively low level, which is the second flat period. According to the established model, the non-linear least squares method is used to fit and analyze the case data in this time period. The initial value is selected as:S(0)= 55699400, E_1_(0)= 150000, E_2_(0)=3000, I_1_(0)=3784, I_2_(0)=3000, A(0)=3000. The fitting result is shown in Fig.8. It can be seen that the fitting result is very good. Some parameters obtained by fitting are shown in Table.2. The proportion of the number of symptomatic infections tested to the total number of symptomatic infections is ρ=0.0056, which means that during this period in England, only 0.56% of symptomatic infections participated in the test. The recovery rates of symptomatic and asymptomatic infected persons non-tested are γ_2_=0.1 and γ_3_=0.35, respectively. It can be seen that the recovery period of symptomatic and asymptomatic infected persons non-tested is about 10 days and 3 days, respectively. The conversion rate from pre-exposure to asymptomatic infection is ε=0.6703, which indicates that it takes about 2 days for a patient in the pre-exposure period to develop into asymptomatic infection.

**Fig.8.**
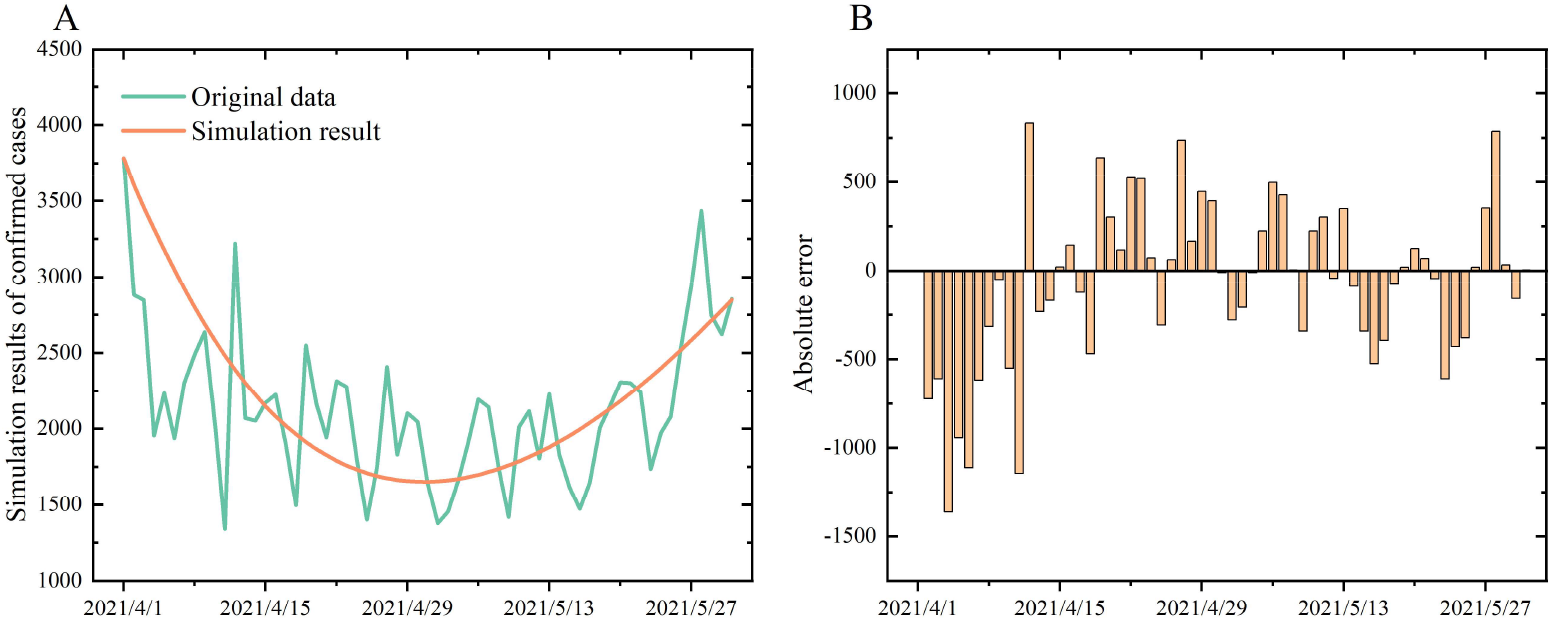
A represents the simulation result of the number of newly confirmed cases per day during the second flat period. B represents the absolute error between the real data and the simulation result.

**Tab.2.** Fitted parameter results during the second flat period

| Parameter | Definition | Value | Source |
| --- | --- | --- | --- |
| $\beta$ | basic transmission rate | $5.7152 \cdot 10^{-9}$ | [21] |
| $\sigma$ | conversion rate of patients in the pre-exposure period to patients in the post-exposure period | $1/4.4$ | [20] |
| $\varepsilon$ | conversion rate from patients in the pre-exposure period to asymptomatic infections | 0.6703 | Fitted |
| $\alpha$ | conversion rate from patients in the post-exposure period to symptomatic infections | 1/2 | [13] |
| $\omega$ | transmission intensity of asymptomatic infection | 0.6524 | [7] |
| $\rho$ | rate of symptomatically infections tested | 0.0056 | Fitted |
| $\gamma_1$ | recovery rate of symptomatic infections tested | 1/17 | [14-16] |
| $\gamma_2$ | recovery rate of symptomatic infections non-tested | 0.1000 | Fitted |
| $\gamma_3$ | recovery rate of asymptomatic infections | 0.3547 | Fitted |
| $\varphi_1$ | mortality rate of symptomatic infections tested | $1.7826 \times 10^{-5}$ | [17] |
| $\varphi_2$ | mortality rate of symptomatic infections non-tested | 0.0010 | Fitted |

Re-predicted according to the parameters obtained by the fitting, and the result is shown in Fig.9B. It can be seen that the number of new confirmed cases per day began to rise from around May 7, 2021, and after about 73 days of development of the epidemic, it reached a peak around July 20. In terms of the overall trend of the development of the epidemic, the predicted results of the model are consistent with the actual occurrence of cases. It can be seen from Fig.9 that from the comparison of the development process of the two epidemics, the development process of the epidemic after the second flat period has been shortened by nearly two months, which also confirms that the Delta variant is more infectious from the perspective of model fitting.

**Fig.9.**
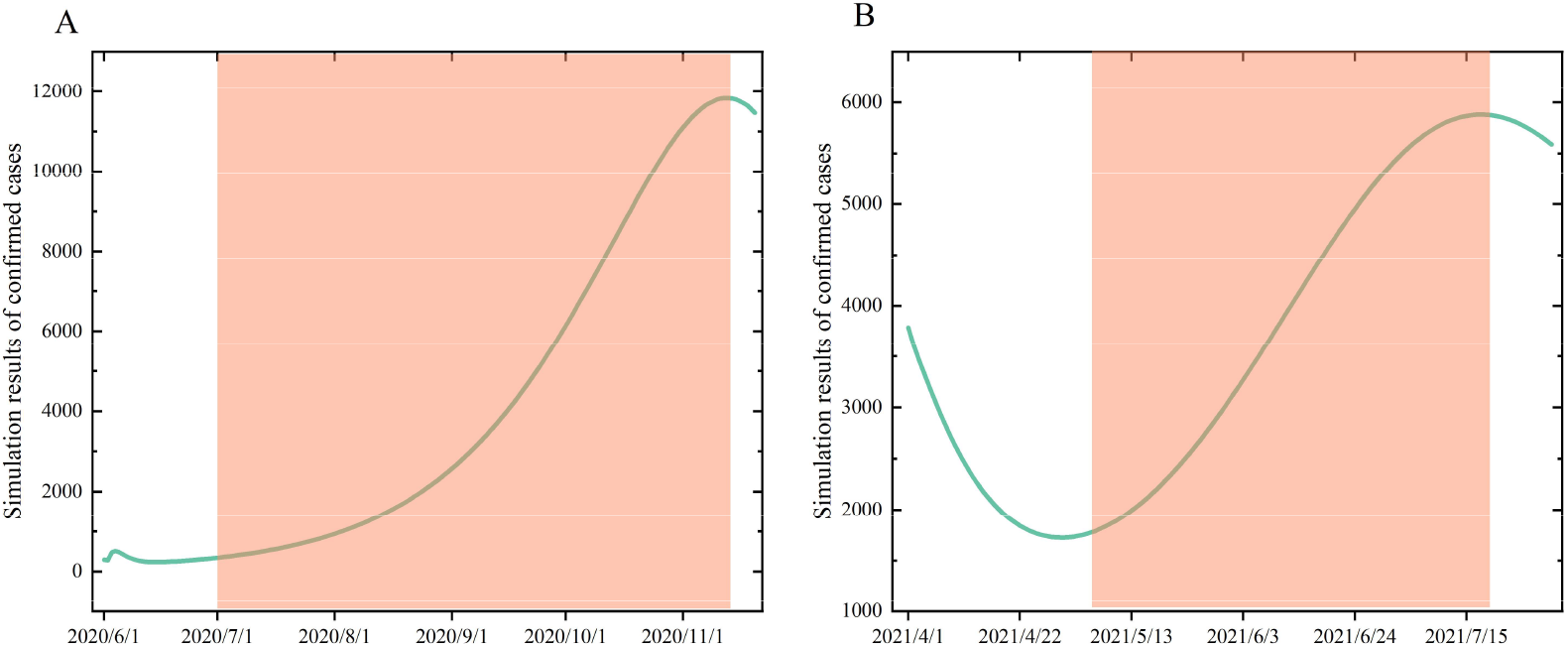
A represents the simulation result of the number of confirmed cases per day after the first flat period. B represents the simulation result of the number of confirmed cases per day after the second flat period. The orange area is the period from the beginning to the peak of the epidemic. It can be seen from the comparison of the two sub-pictures that the epidemic developed more rapidly after the second flat period, and the time was shortened by nearly two months. This also confirms the more infectious characteristics of the Delta variant from the perspective of model simulation.

According to the fitted parameters, the cumulative amount and proportion of infected persons in the second flat period are predicted, as shown in Fig.10. From the predicted results, the proportion of asymptomatic infections rose to 53%.

**Fig.10.**
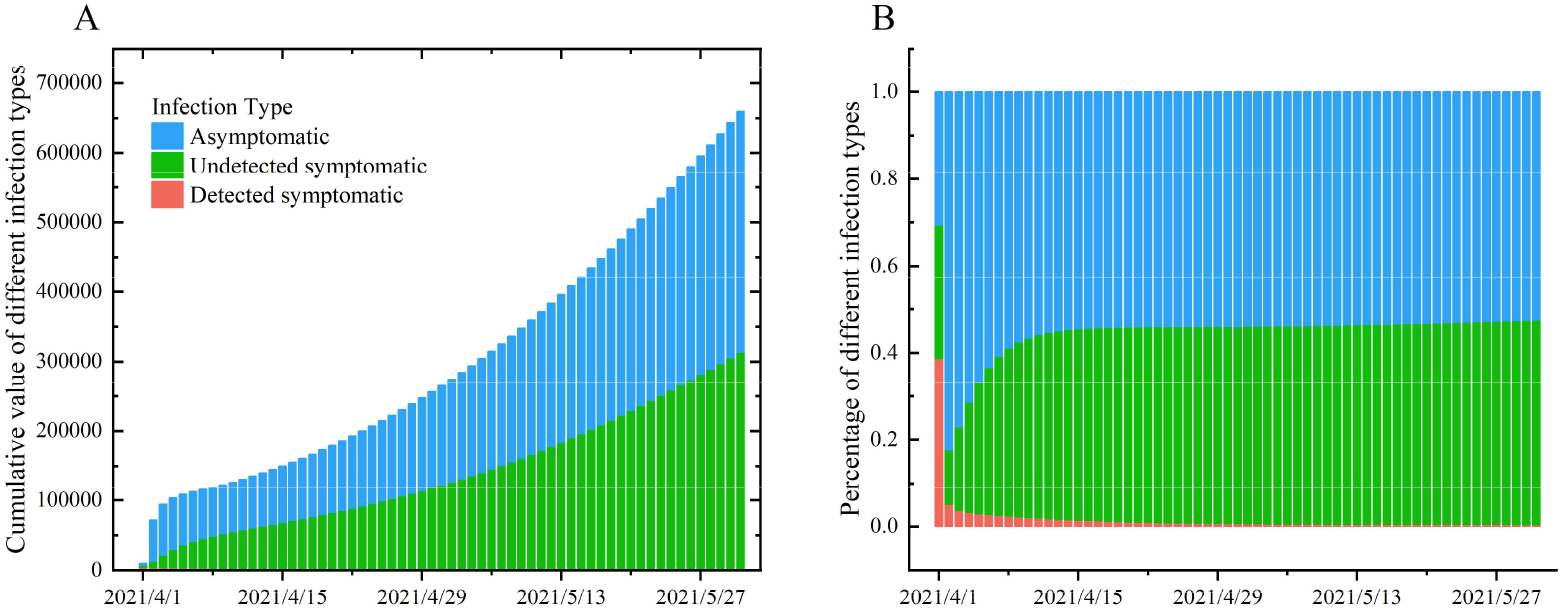
A represents the cumulative value of the simulation results of the three types of infected persons in the second flat period. B represents the proportion of the three types of infections, of which the proportion of asymptomatic infections is about 53%.

### 2.5 Data simulation and analysis in the third flat period

From September 1 to October 18, 2021, the epidemic was at a relatively low level, but the number of new cases per day remained at a relatively high flat period. During this time, the Delta variant was still popular in England. According to the established model, the non-linear least squares method is used to fit and analyze the case data of the third flat period. The initial value is selected as:S(0)=55699400, E_1_(0)= 150000, E_2_(0)=30000, I_1_(0)=30000, I_2_(0)=30000, A(0)=30000. As shown in Fig.11, it can be seen that the fitting results are very good. Some parameters obtained by fitting are shown in Table.3. Fittingly, the proportion of the number of symptomatic infections tested to the total number of symptomatic infections is ρ=0.0779, which means that during this period in England, the proportion of symptomatic infections who participated in the test was only 7.79%. The recovery rates of symptomatic infections non-tested and asymptomatic infections are γ_2_=0.01 and γ_3_=0.3015, respectively, which means that the recovery period of symptomatic infections non-tested is about 10 days, while the recovery period of asymptomatic infections is about 4 days. The conversion rate from pre-exposure to asymptomatic infection is ε=0.7647, which indicates that it takes about 2 days for a patient in the pre-exposure period to develop into asymptomatic infection.

**Tab.3.** Fitted parameter results during the third flat period

| Parameter | Definition | Value | Source |
| --- | --- | --- | --- |
| $\beta$ | basic transmission rate | $5.7152 \times 10^{-9}$ | [21] |
| $\sigma$ | conversion rate of patients in the pre-exposure period to patients in the post-exposure period | 1/4.4 | [20] |
| $\varepsilon$ | conversion rate from patients in the pre-exposure period to asymptomatic infections | 0.7647 | Fitted |
| $\alpha$ | conversion rate from patients in the post-exposure period to symptomatic infections | 1/2 | [13] |
| $\omega$ | transmission intensity of asymptomatic infection | 0.6524 | [7] |
| $\rho$ | rate of symptomatically infections tested | 0.0779 | Fitted |
| $\gamma_1$ | recovery rate of symptomatic infections tested | 1/17 | [14][15][16] |
| $\gamma_2$ | recovery rate of symptomatic infections non-tested | 0.1000 | Fitted |
| $\gamma_3$ | recovery rate of asymptomatic infections | 0.3015 | Fitted |
| $\varphi_1$ | mortality rate of symptomatic infections tested | $1.7826 \times 10^{-5}$ | [17] |
| $\varphi_2$ | mortality rate of symptomatic infections non-tested | 0.0010 | Fitted |

**Fig.11.**
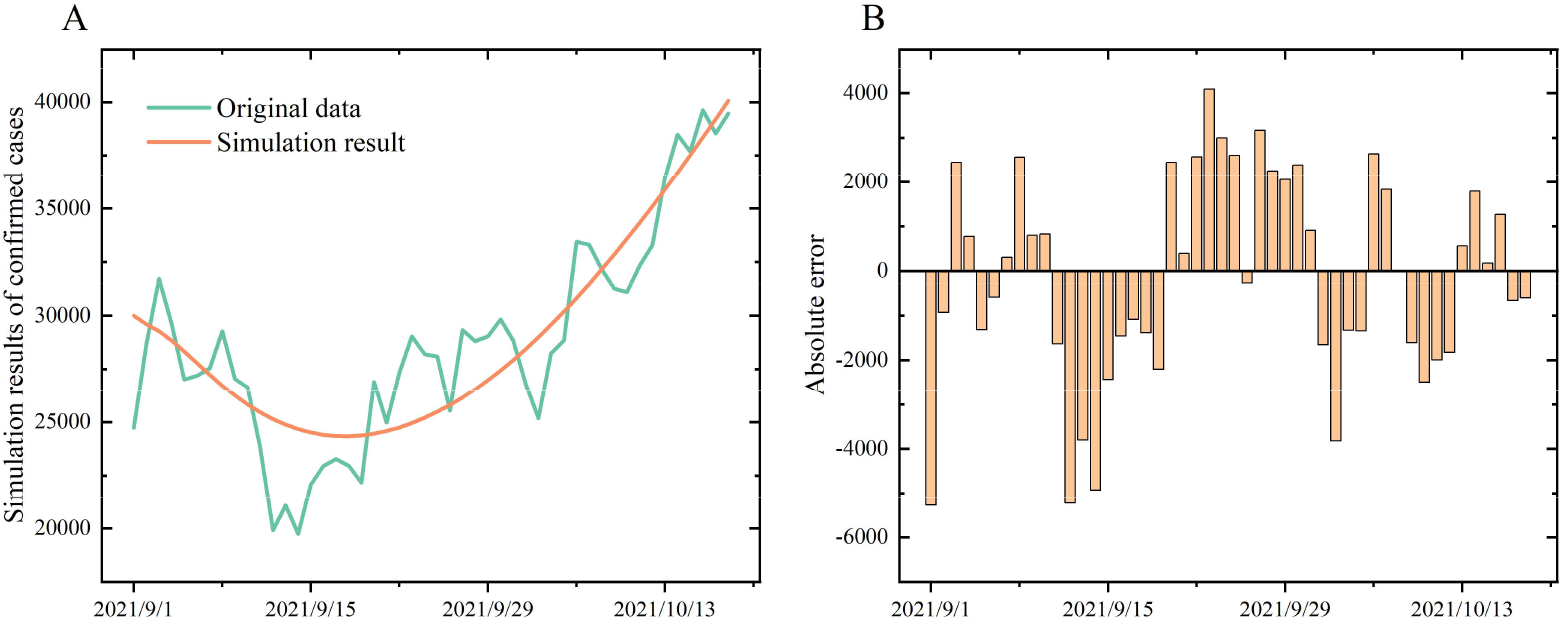
A represents the simulation result of the number of newly confirmed cases per day during the third flat period. B represents the absolute error between the real data and the simulation result.

According to the fitted parameters, the cumulative amount and proportion of infected persons in the second flat period are predicted, as shown in Fig.12. From the predicted results, the proportion of asymptomatic infections rose to 58%. Compared with the fitting results of the first two flat periods, the proportion of asymptomatic patients in the third flat period continued to rise.

**Fig.12.**
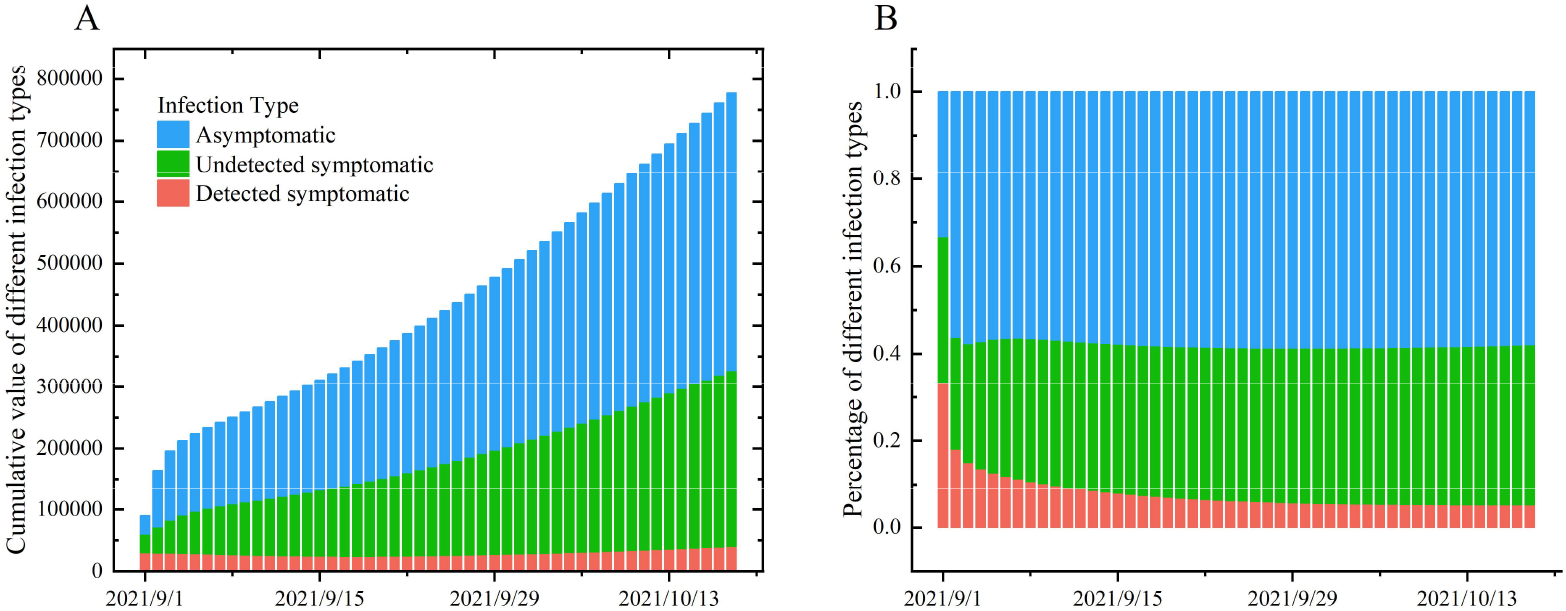
A represents the cumulative value of the simulation results of the three types of infected persons in the third flat period. B represents the proportion of the three types of infections, of which the proportion of asymptomatic infections is about 58%.

Based on the parameters obtained by the previous fitting, the development of the epidemic after the third flat period was predicted. The results are shown in Fig.13. It can be seen that this round of epidemics began to rise in October 2021, reached a peak around early December 2021, then began to decline, and will eventually continue until May 2022.

**Fig.13.**
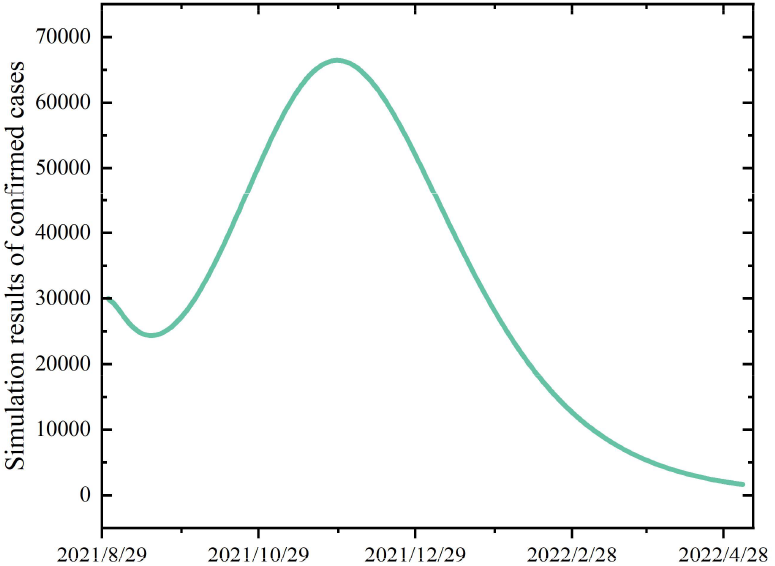
This figure represents the model simulation results of daily newly confirmed cases after the third flat period. As can be seen from the figure, the current round of the epidemic will reach its peak in early December and will continue until May 2022.

## 3 Conclusion

It is difficult for asymptomatic infections to be actively detected, which makes it difficult to cut off the transmission of asymptomatic infections under relatively loose epidemic prevention policies. Especially after the last round of the epidemic is over, the epidemic prevention policy will often be gradually liberalized as the number of confirmed cases per day drops, which will cause the number of asymptomatic infections to accumulate again. At the same time, the number of daily diagnoses has gradually decreased and remained at a low level, which will instead make misjudgments about the current epidemic situation. This is also the reason why the outbreaks in many countries continue to repeat. Therefore, the analysis and evaluation of asymptomatic infections are also very important.

This article established a COVID-19 transmission dynamics model including asymptomatic infections, and discussed the role of asymptomatic infections in the early stages of the spread of the epidemic. Based on the established model, we fitted the epidemic data in the three time periods in England and analyzed the development of the epidemic in the three time periods. During the first flat period, the proportion of people with symptomatic infection tested was ρ=39.55%, and the conversion rate from patients in the pre-exposure period to asymptomatic infections was ε=0.1473. During the second flat period, the proportion of people with symptomatic infection tested was ρ=0.56%, and the conversion rate from patients in the pre-exposure period to asymptomatic infections was ε=0.6703. During the first flat period, the proportion of people with symptomatic infection tested was ρ=7.79%, and the conversion rate from patients in the pre-exposure period to asymptomatic infections was ε=0.7647. According to the obtained parameters, according to the calculation of the model, the proportion of asymptomatic infections in these three flat periods are 41%, 53% and 58% respectively. After the first flat period, the number of daily newly confirmed cases predicted by the model began to increase around July 1, 2020. After more than four months of epidemic spread, it reached a peak on November 12, which is consistent with the actual case situation. Unanimous. After the second flat period, the model predicts that the number of new confirmed cases per day will increase from about May 7, 2021, and after about 73 days of epidemic development, it will reach a peak on July 20, showing the overall trend of the epidemic. In the above, the predicted results of the model are consistent with the actual cases. After the third flat period, the number of daily newly diagnosed cases predicted by the model began to increase around December 1, 2021, and reached a peak in December, and the number of cases will drop to a very low level after May 2022.

## 4. Discussion

SARS-CoV-2 has spread worldwide for almost 2 years. After many mutations, the Delta variant has become the main epidemic virus strain in the world. As we are about to end this part of the research work, Omicron variant has appeared, and infected persons with Omicron variant have been found in many countries and regions. From our research results, because of the large number of asymptomatic infections, the spread of the epidemic is not easy to be completely stopped in a short period of time. However, when the epidemic is in a flat period, nucleic acid testing and quarantine for asymptomatic infected persons at home for 14 days may be an option that can be considered to stop the spread of the epidemic.

Large-scale social activities will also cause the spread of the epidemic in the short term. Taking the Tokyo Olympics as an example, not only did the city where the competition was held, the epidemic situation occurred, but the same phenomenon also appeared in the cities where the competition was not held. During the Olympic Games, Japan’s anti-epidemic policy was relaxed and social mobility increased. This caused a large number of asymptomatic infections to accumulate in the population, which led to a full-scale outbreak of the epidemic. For another example, the beginning of school is also a large-scale social activity. And the campus environment is more closed, which will accelerate the spread of the epidemic on campus. A German study on the transmission of asymptomatic infections among children showed that only 0.4% of asymptomatic children tested positive before admission ^[22]^. In other words, the number of asymptomatic infections among children is likely to be seriously underestimated.

Nucleic acid testing is still one of the most direct and effective methods to control the spread of the epidemic. In many countries and regions, there have been researches on the detection ratio ^[23,24,25]^. Due to factors such as the epidemic situation, anti-epidemic policies, and testing efforts of various countries, there are certain differences in the research results. The existing vaccines, including Pfizer vaccine and Moderna vaccine, still have a certain protective effect against the Delta mutant strain ^[28]^. Vaccination has been around for nearly a year, but the worldwide epidemic still shows no signs of termination or weakening. Repeated lockdown measures and repeated epidemics have not only brought huge economic losses, but also brought more panic and mental exhaustion to people’s daily lives. We will consider and evaluate the role of vaccines in our next work.

## Data Availability

All data produced in the present work are contained in the manuscript；
All data produced are available online

## Acknowledgments

This study was funded by Natural Science Foundation of China (NSFC 11871093), Postgraduate Teaching Research and Quality Improvement Project of BUCEA (J2021010), BUCEA Post Graduate Innovation Project (2021098, 2021099).We thank all the individuals who generously shared their time and materials for this study.

## Notes

### Competing Interest Statement

The authors have declared no competing interest.

